# Reproductive outcomes in patients with uterine fibroid and infertility after laparoscopic myomectomy

**DOI:** 10.1101/2024.05.30.24308110

**Authors:** Nikolai Ruhliada

## Abstract

Uterine fibroids are one of the most common tumors in women worldwide. Considering the negative impact of uterine fibroids on pregnancy in women of reproductive age, myomectomy is the operation of choice. We examined reproductive outcomes in patients undergoing laparoscopic myomectomy for various types of fibroid nodules.

**Objective:** To evaluate reproductive outcomes in patients with infertility and uterine fibroids after laparoscopic myomectomy.

**Materials and methods:** The study included 38 women aged 18 to 45 years with uterine fibroids and infertility. All patients underwent elective laparoscopic myomectomy. The study was conducted among patients who applied routinely to the St. Luke’s Clinical Hospital of St. Petersburg in 2021. At least 2 years of follow-up after possible surgical treatment was acceptable as a time interval for conception. Pregnancy was confirmed by visualizing the fertilized egg in the uterine cavity. Data on the type and number of laparoscopic myomectomies and the characteristics of myoma nodes, such as their number, size and location, were collected from medical records. Obstetric and gynecological history data was also obtained, including the number and outcomes of pregnancies.

**Results:** The study found that of 38 patients with uterine fibroids and infertility, 24 women (63%) became pregnant within 2 years after laparoscopic myomectomy. Of these, 13 (54.1%) were delivered by cesarean section, and 11 (45.9%) were delivered naturally. Of the 5 women with subserous fibroid nodes (FIGO type 7), 5 (100%) became pregnant. Of the 19 patients with intramural subserous nodes (FIGO types 5 and 6), 11 (58%) became pregnant. Of the 24 women with intramural nodes (FIGO type 4), 8 (33%) became pregnant. In 5 (100%) women who became pregnant after removal of the subserous node (FIGO type 7), vaginal delivery was performed. In 7 (63.6%) patients who became pregnant after removal of an intramural-subserous node (FIGO types 5 and 6), delivery was performed by cesarean section, in 4 (36.4%) by natural delivery. In 6 (75%) women who became pregnant after removal of an intramural node (FIGO type 4), delivery was performed by cesarean section, in 2 (35%) by natural delivery. Of 14 women suffering from infertility and having only 1 fibroid node, 11 became pregnant (78.5%), of 19 patients with 2 fibroids, 11 (58%) became pregnant, of 5 women who had 3 or more fibroid nodes became pregnant 2 (40%). Of the 11 women who became pregnant after myomectomy of 1 node, 5 (45.4%) had a cesarean section and 6 (54.6%) had a natural delivery. Of the 11 women who became pregnant after removal of 2 fibroids, 6 (54.5%) had a cesarean section, 5 (45.5%) had a vaginal delivery, of 2 women who became pregnant after removal of 3 or more fibroids, 2 (100%) were performed by caesarean section. In addition, we found that out of 4 women who had fibroids measuring less than 3 cm, only 1 (25%) became pregnant; out of 9 patients with fibroids from 3 to 5 cm, 7 (29%) became pregnant; out of 25 patients with 16 (67%) became pregnant with fibroids larger than 5 cm. In the only woman who became pregnant after myomectomy of a node less than 3 cm, delivery was performed by cesarean section. 4 (57%) patients who became pregnant after removal of fibroids measuring 3 to 5 cm had a cesarean section delivery, and 3 (43%) had a natural delivery. In 8 (50%) patients who became pregnant after removal of a node measuring more than 5 cm, delivery was performed by cesarean section, in 8 (50%) by natural delivery.

**Conclusions:** Depending on the size, number and location, uterine fibroids can cause infertility. Our study demonstrates that laparoscopic myomectomy improves reproductive outcomes in women with subserosal, intramural-subserosal, and intramural myomatous nodules. After removal of nodes larger than 5 cm, the pregnancy rate was significantly higher than after removal of nodes smaller than 5 cm. In addition, the pregnancy rate in this observation period was higher in patients after removal of a single uterine fibroid, compared with women with 2 and more myomatous nodes. After removal of the subserous nodes, delivery was carried out naturally; after removal of the intramural and subserosal-intramural nodes, in most cases the tactics of delivery by cesarean section were chosen. In the groups of women who became pregnant after removal of a node from 3 to 5 cm and more than 5 cm, the rate of delivery by cesarean section and vaginally was approximately equal. After myomectomy of a node less than 3 cm, delivery was carried out naturally. In the groups of women who became pregnant after removal of 1 and 2 nodes, the rate of surgical and natural delivery was approximately the same, and in the group of patients with 3 or more nodes, a cesarean section was performed in all cases.

## Introduction

Uterine fibroids are a benign monoclonal tumor arising from smooth muscle cells of the cervix or uterine body [50]. The incidence of the disease among women of reproductive age reaches 70% [2,34,36]. Most women are asymptomatic, making it difficult to estimate the true prevalence. Symptoms and their severity may vary depending on the size and location of the fibroids. The most common symptom is abnormal uterine bleeding, which can lead to anemia. Other possible symptoms include lower abdominal pain of varying nature and intensity, painful and heavy menstruation, dyspareunia, dysfunction of adjacent organs (dysuric disorders, constipation) [21]. Risk factors for the development of fibroids include early menarche, no history of childbirth, increased BMI, smoking, and vitamin D deficiency [16,42,50]. The average age of detection of uterine fibroids is 32-34 years [36]. Currently, there is an increase in the incidence of fibroids in young women under 30 years of age who have not achieved reproductive function. Depending on their size, number and location in the uterus, fibroids can cause both infertility and recurrent miscarriage [30,46,49]. The location of fibroids in the uterus can be classified using the International Federation of Gynecology and Obstetrics (FIGO) system. In women who need fertility preservation, fibroids can be removed surgically (myomectomy) by laparotomy, laparoscopy, or hysteroscopy, depending on the size, location, and type of fibroids. However, myomectomy is an operation that is not without risks and can lead to serious complications. Therefore, it is important to determine whether myomectomy can lead to improved reproductive outcomes and, if so, to identify the optimal surgical approach.Although there are many studies evaluating the relationship between uterine fibroids and infertility, the mechanisms by which fibroids negatively impact reproductive function remain largely unknown. Many hypotheses have been proposed to explain how fibroids can cause infertility, such as increased uterine contractility, abnormal cytokine profile, abnormal vascularization, and chronic inflammation [19]. The tumor can disrupt the anatomy of the uterine cavity, causing obstruction of the proximal openings of the fallopian tubes and the anatomy of the pelvic organs, pushing the fallopian tube away from the ovary [20]. It is likely that the displacement of the cervix towards the symphysis by large myomatous nodes makes it difficult for sperm to enter it, and an enlargement or deformation of the uterine cavity may interfere with the normal transport of sperm [19]. Myomatous nodes, especially with a submucosal location, disrupt blood flow in the endometrium and cause an excessive inflammatory reaction, which negatively affects implantation [17]. Patients with fibroids also have low levels of interleukin 10 and glycodelin. These cytokines are involved in the implantation process and their decrease can have a negative impact on conception [22].

It is known that fibroids are more common in women with a history of infertility. Fibroids may be the only cause of infertility in 2-3% of women. It is also found in up to 23% of cases of primary and secondary infertility. This is also supported by a recent large cohort study that showed an increase in time to first and subsequent pregnancies in patients with uterine fibroids compared with controls without fibroids [18]. In some cases, uterine fibroids do not prevent pregnancy and fetal development, but increase the incidence of complications of pregnancy and childbirth [17,30,46,49].

Although there are many studies evaluating the relationship between uterine fibroids and infertility, the mechanisms by which fibroids negatively impact reproductive function remain largely unknown. Many hypotheses have been proposed to explain how fibroids may cause infertility, such as increased uterine contractility, abnormal cytokine profiles, abnormal vascularization, and chronic inflammation. The tumor can disrupt the anatomy of the uterine cavity, causing obstruction of the proximal openings of the fallopian tubes and the anatomy of the pelvic organs, pushing the fallopian tube away from the ovary [27]. It is likely that the displacement of the cervix towards the symphysis by large myomatous nodes makes it difficult for sperm to enter it, and an enlargement or deformation of the uterine cavity may interfere with the normal transport of sperm. Myomatous nodes, especially with a submucosal location, disrupt blood flow in the endometrium and cause an excessive inflammatory reaction, which negatively affects implantation [17]. The negative effects of intramural fibroids may also be mediated by signaling molecules produced by the fibroids that are able to reach the endometrial cavity, causing adverse effects on endometrial homeostasis and receptivity [17,49].

The location of the fibroid nodes obviously determines the degree of influence on the reproductive outcome. Previous studies have shown that women with submucosal fibroids that deform the uterine cavity, compared with women without fibroids, have a higher incidence of infertility, miscarriage, placental insufficiency, placenta previa, premature placental abruption and fetal malposition, and a lower incidence of implantation and childbirth The incidence of cesarean section and preterm birth is also likely to increase in the presence of submucosal fibroids [6,12]. On the other hand, subserosal fibroids do not have a significant impact on reproductive outcome, while the impact of intramural fibroids remains controversial due to conflicting study results [33,37,40].

There is evidence that fibroid size and proximity to the uterine cavity are important for the effect of intramural fibroids on fertility. Accordingly, fibroids as small as 2 cm located close to the endometrial mucosa (FIGO type 3) will have a detrimental effect on reproductive outcome. In the case of intramural fibroids that are not in contact with the underlying endometrium (type 4, 5), 3 cm is the minimum size that negatively affects a woman’s fertility [8]. Women with fibroids may also experience higher rates of fetal growth restriction and stillbirth [23]. However, a recent large study of more than 5,500 women found no statistically significant increase in the risk of spontaneous abortion among women with and without fibroids [15].

The effect of fibroid number on fertility has not been well studied, although fewer fibroids have been reported to be associated with improved reproductive outcomes after fibroid removal [29]. However, some studies have not found a relationship between fibroid size and pregnancy rates [20].

Studies comparing the results of IVF cycles in women with intramural fibroids and women without fibroids demonstrate a significant negative impact of intramural fibroids on fertility and recommend surgical removal of fibroids before IVF [1,11,32,33,45,47]. Studies confirm that surgery is indicated for intramural fibroids affecting the cavity, but not for subserous fibroids. Intramural nodes larger than 4 cm in size that do not deform the uterine cavity are recommended to be removed before IVF. On the other hand, the role in the genesis of infertility and the benefits of myomectomy on reproductive outcomes in intramural fibroids (FIGO) types 3–5 are less clear [13,25]. However, some authors were unable to detect a negative effect on IVF outcomes in the presence of fibroids less than 5 cm in size and without disruption of the structure of the uterine cavity [3,35].

There is currently insufficient evidence that the specific size, number or location of fibroids (excluding submucosal fibroids or intramural fibroids affecting the structure of the uterine cavity) is associated with a decreased likelihood of pregnancy or an increased risk of early pregnancy loss [32].

The choice of management tactics for patients with infertility and intramural or subserous fibroids depends on the clinical symptoms, as well as the size and location of the fibroids. If significant symptoms associated with fibroids occur, such as heavy menstrual bleeding or severe pain, fibroid removal is usually recommended to relieve symptoms. However, when women experience infertility or recurrent miscarriage in the absence of fibroid-related symptoms, treatment recommendations are less clear.

Drug treatment may be used in the presence of abnormal uterine bleeding, although this approach has no more than a temporary effect on fibroids. In certain cases, GnRH agonists can be used preoperatively to reduce fibroid size and restore hemoglobin levels in symptomatic patients [14,26].

Myomectomy is the leading method of restoring fertility in patients with infertility or miscarriage. Studies have shown that myomectomy significantly increases pregnancy rates in women with fibroid-related infertility. The incidence of miscarriage decreases after myomectomy, although overall rates of spontaneous pregnancy loss remain higher than in the general population. Fibroids affect 0.1– 3.9% of pregnant women, and a number of complications occurring during the prenatal and postpartum period are believed to be directly related to the presence of these benign tumors [17]. The positive effect of myomectomy on reproductive prognosis is also proven by a study in which 52.2% of women became pregnant after myomectomy [17]. In another study, women who underwent myomectomy had higher birth rates compared with women with fibroids who did not undergo surgery (42% versus 11%, respectively) [32].

Myomectomy improved reproductive outcome in women with fibroids that distort the uterine cavity [48]. Improved reproductive outcomes after myomectomy were also observed in a group of women who had at least one fibroid with a diameter of more than 5 cm that did not distort the uterine cavity [13]. Myomectomy was not associated with improved pregnancy rates in women with intramural or subserous fibroids smaller than 4 cm [26]. Most scientists agree that subserous fibroids do not affect the incidence of implantation, clinical pregnancy, childbirth and early pregnancy termination, and its removal is not beneficial in the absence of special indications for surgical treatment (torsion of the node leg, poor circulation in the node, symptoms compression of adjacent organs, rapid growth of the node) [31,32].

Some authors report no clear benefit from surgery and do not recommend this approach. However, a limitation of these studies is that they do not provide clear information on the size, number and location of fibroids [7,23,26].

Don E et al. (2022) compared reproductive outcomes in patients after myomectomy and patients with fibroids. The study found no significant difference in time to live birth after myomectomy compared with expectant management in women with fibroids who want to become pregnant. However, the majority of conceptions in the myomectomy group occurred naturally (67%), while IVF/ICSI (45%) was the most common method of conception in the fibroid group. After a woman has had a myomectomy, she has a recovery period of up to 6 months, during which she is advised not to become pregnant. Therefore, the recovery period after surgery may have been offset by the possible beneficial effect of myomectomy, which, in addition to the reduction of symptoms, provides an argument in favor of performing myomectomy.

Most authors concluded that there is insufficient evidence regarding the effect of removal of intramural fibroids on reproductive outcomes in infertile women [32].

Intramural and subserous myomatous nodes can be removed using laparoscopic, laparotomic access, as well as by performing a mini-laparotomy. Laparoscopic myomectomy is the “gold standard” in the absence of contraindications. It has some advantages over laparotomy: less postoperative pain, shorter hospitalization time, less blood loss and faster recovery [24,27].

In a study conducted by Morales H. et al. (2022) the pregnancy rate was statistically significantly higher with laparoscopic surgery compared with laparotomy and expectant management. Tsiampa E et al. (2023) proved that laparoscopic myomectomy provides better results compared to minilaparotomy.

Some studies have reported no differences between laparoscopic and laparotomic approaches in terms of pregnancy rates and early pregnancy loss rates [9,19,26].

The most common intraoperative complications of laparoscopic myomectomy include myometrial hematoma, excessive blood loss, and accidental morcellation [39]. Contraindications to laparoscopic myomectomy are multiple fibroids (> 4) in different parts of the uterus, requiring multiple incisions, and the presence of intramural fibroids larger than 10-12 cm or suspected leiomyosarcoma [9].

The laparotomy approach is preferable in patients with multiple fibroids, as it reduces operative time [38].

Unfortunately, myomectomy is a fairly adhesive surgical procedure, and postoperative adhesions can have a significant impact on fertility [25]. Myomectomy itself, being a serious invasive procedure, carries a risk of damage to the uterine myometrium and endometrium, as well as the formation of adhesions in the pelvic cavity. Additionally, the direct cause-and-effect relationship between the presence of fibroids and infertility and the actual benefit of myomectomy has not yet been determined. Thus, in women with unexplained infertility or requiring treatment of symptomatic fibroids, the surgeon must balance the benefits of such surgery in terms of improving fertility on the one hand, and the consequences associated with the development of adhesions, on the other, while avoiding unnecessary myomectomies and involuntary iatrogenic injuries. This approach is especially relevant in the case of large fibroids located on the posterior wall. It is therefore important to determine when myomectomy should be considered beneficial and, if so, to take all available measures to avoid postoperative infertility [25].

Currently, hysteroscopic myomectomy is the gold standard for surgical treatment of submucosal fibroids (FIGO 0 and 1 fibroids) [28]. In approximately 70% of cases, submucosal uterine fibroids lead to abnormal uterine bleeding, which is the most common indication for hysteroscopic myomectomy [44]. Other indications include infertility, recurrent miscarriage, dysmenorrhea and pelvic pain [22, 28]. Complications during the intervention are rare, but uterine perforation and bleeding are possible [4,12,22].

There is reliable evidence that hysteroscopic myomectomy for submucosal fibroids that deform the uterine cavity improves clinical pregnancy rates and reduces the risk of early pregnancy loss, especially when fibroids are the only cause of infertility [12,32,22].

There is evidence that damage to the pseudocapsule during myomectomy can lead to a decrease in the content of neuropeptides and neurofibers at the site of hysterotomy, impaired physiological healing of the myometrium with increased fibrosis due to hypoxia, ischemia and necrosis. These pathophysiological phenomena cause a deficiency of neurotransmission in the myometrium, muscle impulses and contractility, which ultimately leads to dysfunction of the uterine muscles during pregnancy and childbirth. Tinelli et al. (2019) recommended preservation of the pseudocapsule during myomectomy. Hysteroscopic cold snare myomectomy appears to reduce thermal injury and preserve the integrity of the fibroid pseudocapsule.

## Materials and methods

The study included 63 women aged 18 to 45 years at the time of diagnosis of uterine fibroids. All patients underwent laparoscopic myomectomy. The study was conducted among patients who applied to the St. Luke’s Clinical Hospital of St. Petersburg as planned in 2021. At least 2 years of follow-up after possible surgical treatment was acceptable as a time interval for conception. Patients with severe endometriosis (grade 4) or dominant adenomyosis (as adenomyosis affects the uterus to a greater extent than uterine fibroids) were excluded.

Data were collected from medical records on the type and number of myomectomies (hysteroscopic/laparoscopic/laparotomy myomectomy), fibroid characteristics such as number, size and location. Obstetric and gynecological history data was also obtained, including the number and outcome of pregnancies, possible complications of pregnancy and childbirth.

Reasons for myomectomy included infertility and/or fibroid-related symptoms.

## Results

The study found that of 38 patients with uterine fibroids and infertility, 24 women (63%) became pregnant within 2 years after laparoscopic myomectomy (Tab. 1). Of these, 13 (54.1%) were delivered by cesarean section, and 11 (45.9%) were delivered naturally. Of 38 women planning pregnancy, 5 (13%) had pedunculated subserous fibroids (FIGO type 7), 19 (50%) had intramural-subserous fibroids (FIGO type 5 or 6), 14 (37%) had intramural (FIGO type 4).

Of the 38 women planning pregnancy, 14 (37%) had a single fibroid, 19 (50%) had 2 fibroids, and 5 (13%) had 3 or more.

When assessing the size of fibroids, it was found that in 4 (10.5%) patients out of 38, the size of fibroids was less than 3 cm, in 9 (23.5%) out of 38 from 3 to 5 cm, in 25 (66%) there was fibroids more than 5 cm.

Of the 5 women with subserous fibroids (FIGO type 7), 5 (100%) became pregnant. After laparoscopic removal of fibroids of this location, patients could plan pregnancy within 3-4 months. Of the 19 patients with intramural subserous nodule (FIGO types 5 and 6), 11 (58%) became pregnant. Of the 24 women with an intramural node (FIGO type 4), 8 (33%) became pregnant. After a woman has had a myomectomy, she has a recovery period of up to a year, during which time she is advised not to become pregnant. This factor could also influence the frequency of conception among patients with this type of fibroid. In 5 (100%) women who became pregnant after removal of the subserous node (FIGO type 7), vaginal delivery was performed. In 7 (63.6%) patients who became pregnant after removal of an intramural-subserous node (FIGO types 5 and 6), delivery was performed by cesarean section, in 4 (36.4%) by natural delivery. In 6 (75%) women who became pregnant after removal of an intramural node (FIGO type 4), delivery was performed by cesarean section, in 2 (35%) by natural delivery.

Of 14 women suffering from infertility and having a single fibroid, 11 became pregnant (78.5%), of 19 patients with 2 fibroids, 11 (58%) became pregnant, of 5 women who had more than 3 fibroids, 2 became pregnant (40%). Of the 11 women who became pregnant after myomectomy of 1 node, 5 (45.4%) had a cesarean section and 6 (54.6%) had a natural delivery. Of the 11 women who became pregnant after the removal of 2 fibroids, 6 (54.5%) had a cesarean section, 5 (45.5%) had a natural delivery, of 2 women who became pregnant after the removal of 3 or more fibroids, 2 (100%) were performed by caesarean section.

In addition, we found that out of 4 women who had fibroids measuring less than 3 cm, only 1 (25%) became pregnant; out of 9 patients with fibroids from 3 to 5 cm, 7 (29%) became pregnant; out of 25 patients with 16 (67%) became pregnant with fibroids larger than 5 cm. In the only woman who became pregnant after myomectomy of a node less than 3 cm, delivery was performed by cesarean section. 4 (57%) patients who became pregnant after removal of fibroids measuring 3 to 5 cm had a cesarean section delivery, and 3 (43%) had a natural delivery. In 8 (50%) patients who became pregnant after removal of a node measuring more than 5 cm, delivery was performed by cesarean section, in 8 (50%) by natural delivery. The delivery tactics were determined by the obstetrician of the maternity hospital. In Table 2 we described the distribution by size of uterine fibroids.

**Table 1.**
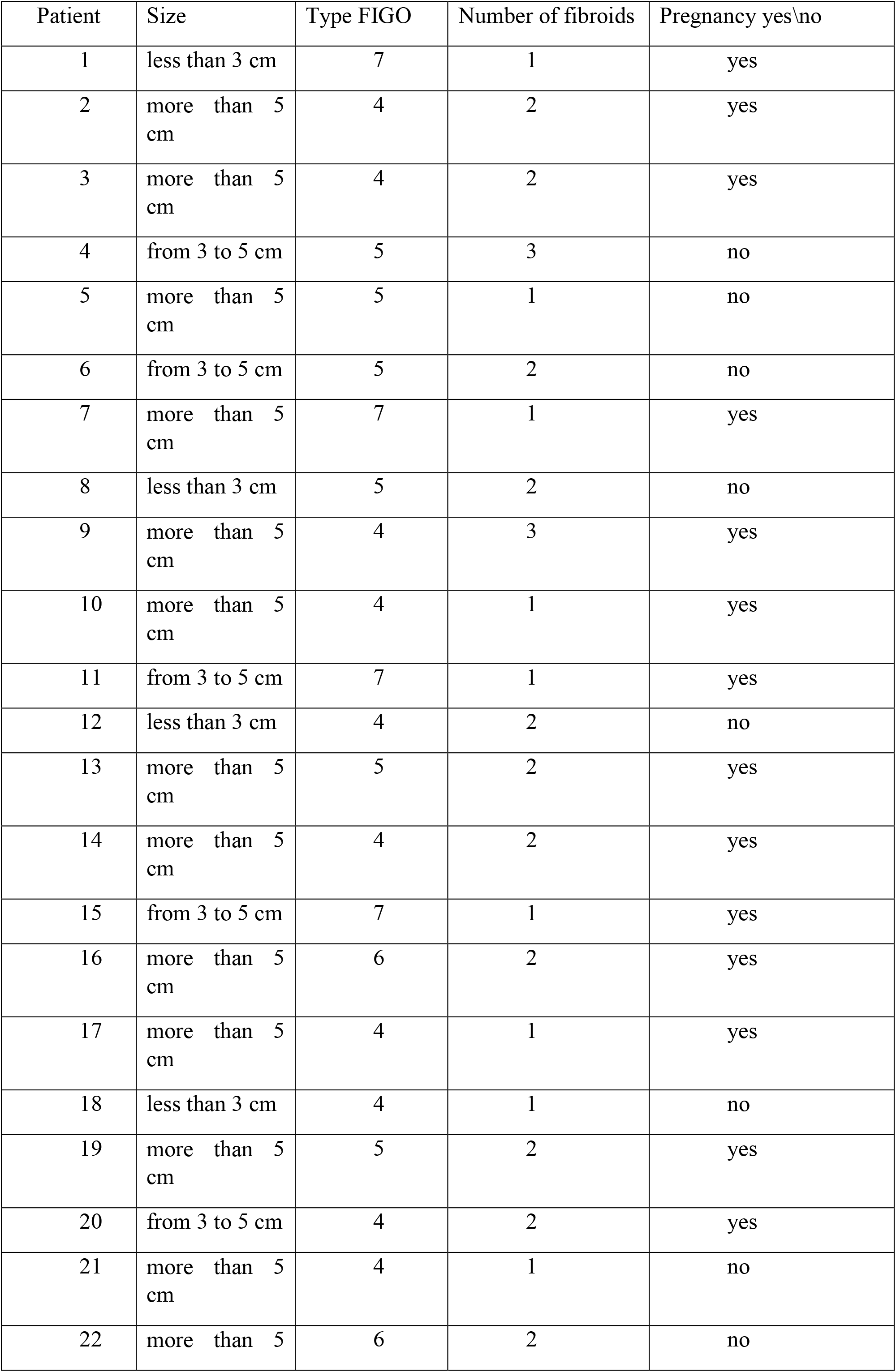
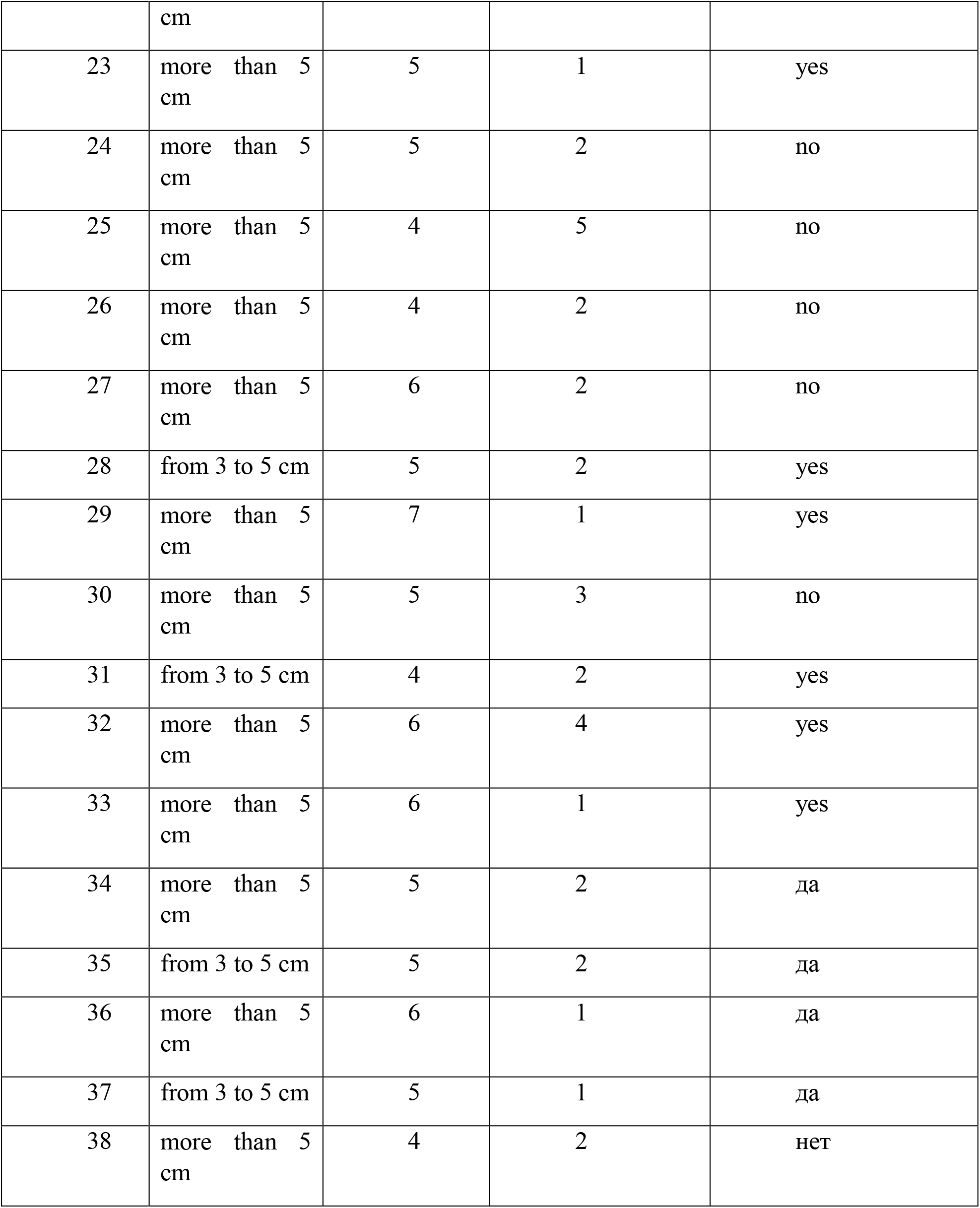
Distribution of patients by type, number of uterine fibroids and reproductive outcomes

**Table 2.** The distribution by size of uterine fibroids

| Patient | size in mm |
| --- | --- |
| 1 | 23x25x20 |
| 2 | 62x74x70 и 55x60x64 |
| 3 | 102x91x95 и 72x70x75 |
| 4 | 38x47x48 и 31x42x45 и 40x43x46 |
| 5 | 55x58x60 |
| 6 | 44x48x45 и 40x46x49 |
| 7 | 51x60x74 |
| 8 | 13x9x11 и 15x20x24 |
| 9 | 85x87x71 и 62x63x74 и 49x51x53 |
| 10 | 87x89x92 |
| 11 | 30x35x42 |
| 12 | 8x8x10 и 20x26x28 |
| 13 | 93x66x64 и 62x93x75 |
| 14 | 77x79x85 и 90x93x95 |
| 15 | 34x38x45 |
| 16 | 88x85x83 и 74x71x68 |
| 17 | 51x64x72 |
| 18 | 10x15x17 |
| 19 | 76x71x75 и 65x60x71 |
| 20 | 33x37x39 и 41x45x48 |
| 21 | 65x69x71 и 86x89x94 |
| 22 | 50x53x55 и 77x74x75 |
| 23 | 51x53x55 |
| 24 | 59x58x54 |
| 25 | 90x94x97 и 54x58x57 и 50x51x54 и 35x30x31 и 45x35x50 |
| 26 | 64x68x69 и 71x75x76 |
| 27 | 54x58x59 и 60x62x65 |
| 28 | 41x45x47 и 31x36x43 |
| 29 | 66x70x72 |
| 30 | 51x60x58 и 66x68x74 и 70x75x76 |
| 31 | 45x40x39 и 31x35x34 |
| 32 | 54x57x62 и 50x53x54 и 90x84x86 и 47x52x53 |
| 33 | 69x75x79 |
| 34 | 70x62x65 и 77x74x80 |
| 35 | 44x47x48 и 30x35x37 |
| 36 | 55x59x62 |
| 37 | 30x34x40 |
| 38 | 95x93x90 и 85x89x86 |

## Conclusions

Depending on the size, number and location, uterine fibroids can cause infertility. Our study demonstrates that laparoscopic myomectomy improves reproductive outcomes in women with subserosal, intramural-subserosal, and intramural myomatous nodules. In addition to the problem of infertility, potential symptoms such as abnormal uterine bleeding and pain should be assessed and included in the indications for surgery. After removal of nodes larger than 5 cm, the pregnancy rate was significantly higher than after removal of nodes smaller than 5 cm. In addition, the pregnancy rate in this observation period was higher in patients after removal of a single uterine fibroid, compared with women with 2 and more myomatous nodes. After removal of the subserous nodes, delivery was carried out naturally; after removal of the intramural and subserosal-intramural nodes, in most cases the tactics of delivery by cesarean section were chosen. In the groups of women who became pregnant after removal of a node from 3 to 5 cm and more than 5 cm, the rate of delivery by cesarean section and vaginally was approximately equal. After myomectomy of a node less than 3 cm, delivery was carried out naturally. In the groups of women who became pregnant after removal of 1 and 2 nodes, the rate of surgical and natural delivery was approximately the same, and in the group of patients with 3 or more nodes, a cesarean section was performed in all cases.

## Ethic committee

Decision 14-h from 12 Feb. 2024. Protocols for research study supervised by professor Ruhliada N.N. were approved (Approval no. 1478/2022) in Compliance with National Institutes of Health guidelines for human studies. Along with that medical records used for this study were anonymized before access.

## Data Availability

All data produced in the present study are available upon reasonable request to the authors

